# Attitudes of the US general public towards Monkeypox

**DOI:** 10.1101/2022.06.20.22276527

**Authors:** Amyn A. Malik, Maike S. Winters, Saad B. Omer

**Author notes:** Corresponding author: Maike S. Winters. Contributed equally (Amyn A. Malik, Maike S. Winters).

## Abstract

While the Monkeypox outbreak is growing, little is known about the general public’s levels of knowledge, their trusted sources of information and attitudes towards a Monkeypox vaccine. In our representative survey of the US general public, we find that almost half the respondents (47%) feel that their knowledge level about Monkeypox is poor or very poor. The most trusted sources of information about the outbreak are healthcare professional and officials, but also known doctors and researchers with a large online following. Being vaccinated against COVID-19 was a strong predictor of willingness to receive a Monkeypox if recommended (adjusted Odds Ratio 32.1, 95% Confidence Interval 16.7-61.7). Our findings point to the urgent need for clear communication.

## Introduction

The World Health Organization has expressed concern about an emerging global outbreak of Monkeypox.^1^ Experience from the COVID-19 pandemic has highlighted the need for early attention to behavioral aspects of a response.^2,3^ However, little is known about the general public’s knowledge, attitudes and practices around Monkeypox and the vaccine against Monkeypox to inform communication campaigns and behavioral interventions. We therefore aimed to survey the US general public about their knowledge and attitudes, their trusted sources of information and to test whether COVID-19 vaccination status was associated with Monkeypox vaccination attitudes.

## Methods

An online survey was administered in the United States. A representative sample of the US was recruited through CloudResearch in early June 2022. Participants answered questions on their awareness and knowledge of Monkeypox, their trusted sources of information, COVID-19 vaccination status, as well as intentions to receive a vaccine against Monkeypox if they would be recommended to do so.

Descriptive statistics were summarized and weighted by age, gender and race (see supplement). Logistic regression analysis was carried out to determine the predictors of intention to take a recommended Monkeypox vaccine. Yale University Institutional Review Board approved this study (IRB protocol number 2000032980) and participants gave their informed consent before data was collected.

## Results

The sample comprised 856 participants, of which 51% was female, 41% had a college degree or higher and 38% was 55 years or older, which was similar to the US population. The sources of information deemed most reliable to convey information about the outbreak were Healthcare Professionals (median on scale from 1-5: 3.7, standard deviation (SD) 1.4), Health Officials (like the Centers for Disease Control and Prevention) (3.5, SD 1.3), and social media accounts of known doctors and researchers (3.1, SD 1.4), Figure 1.

**Figure 1:**
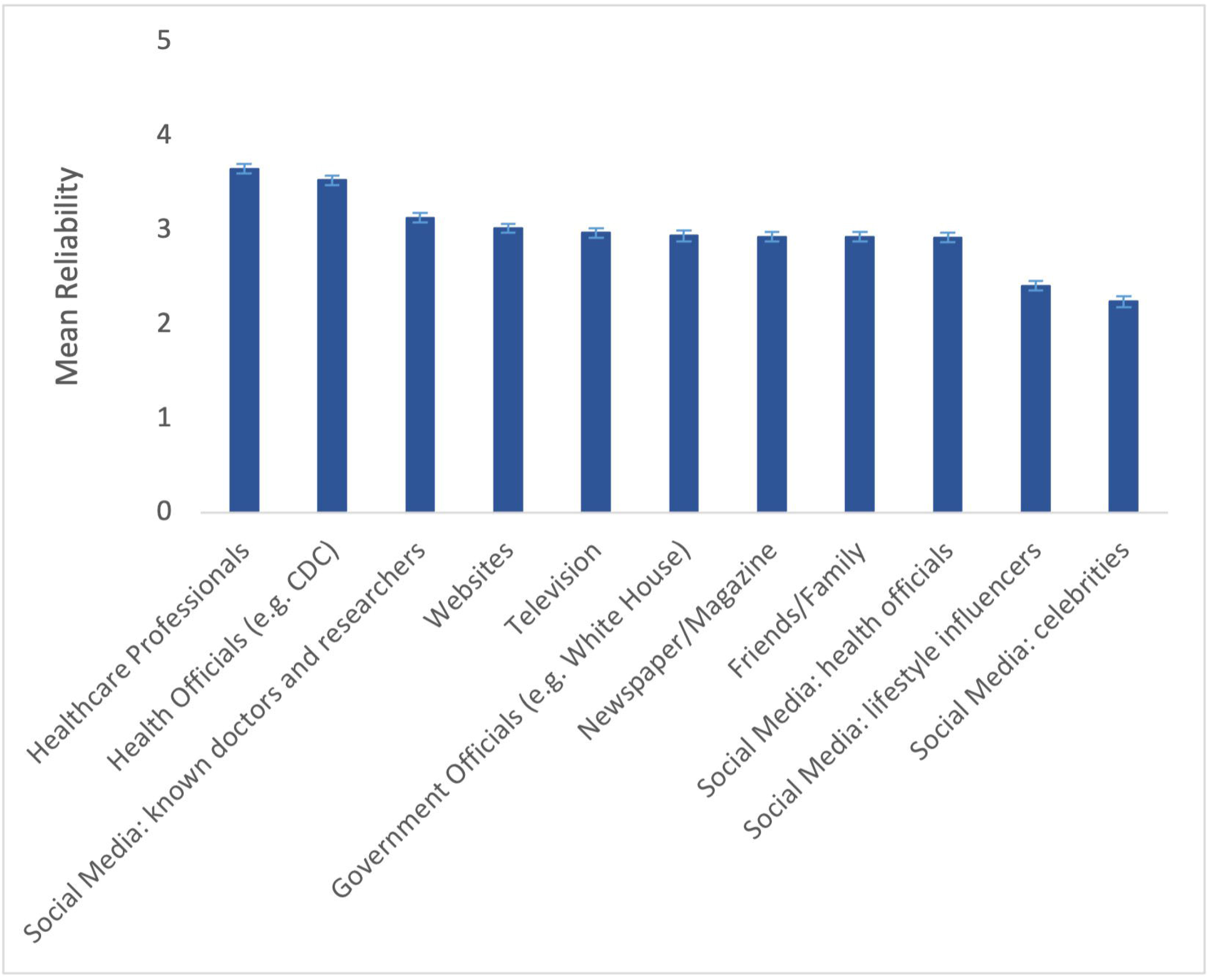
Mean Reliability of Information Sources

When asked about their COVID-19 vaccination status, 68% said they were vaccinated, which matched the share of fully vaccinated individuals in the US (67%).^4^ Of the sample, 79% was aware of the Monkeypox outbreak, but almost half the respondents (47%) rated their level of knowledge about Monkeypox to be poor or very poor. 44% of the respondents were concerned about the outbreak. While most of the respondents pointed to correct preventative measures such as avoiding close contact with sick people (83%) and washing hands with soap and water (80%), 48% said that eating a balanced diet was an effective way to prevent Monkeypox.

Among those older than 45 years (55% of the sample), 67% were vaccinated against smallpox, 19% were not and 14% did not know. Asked whether they would receive a Monkeypox vaccine if recommended, 46% (95% CI: 42%-50%) of our sample said ‘yes’, 29% declined and 25% did not know. Current COVID-19 vaccination status was a strong predictor of this attitude towards a Monkeypox vaccine (adjusted Odds Ratio (aOR) 32.1, 95% Confidence Interval (CI) 16.7-61.7), Table 1. Women were less likely than men to take a recommended Monkeypox vaccine (aOR 0.6, 95% CI 0.4-0.8).

**Table 1:** Associations with intention to receive a Monkeypox vaccine if recommended

|  | **Odds Ratio**<br>**(95% CI)** | **P-value** | **Adjusted Odds**<br>**Ratio\* (95% CI)** | **P-value** |
| --- | --- | --- | --- | --- |
| **Covid-19 vaccination status** |  |  |  |  |
| Not vaccinated | Reference |  | Reference |  |
| Vaccinated | 29.61 (15.68-55.91) | 0.000 | 32.14 (16.73-61.74) | 0.000 |
| **Gender** |  |  |  |  |
| Male | Reference |  | Reference |  |
| Female | 0.42 (0.31-0.58) | 0.000 | 0.56 (0.38-0.83) | 0.004 |
| **Education** |  |  |  |  |
| No high school | Reference |  | Reference |  |
| High school | 1.75 (0.63-4.98) | 0.283 | 0.74 (0.16-3.30) | 0.689 |
| Some college | 2.35 (0.84-6.60) | 0.103 | 0.72 (0.16-3.23) | 0.664 |
| College | 3.69 (1.32-10.28) | 0.013 | 0.77 (0.17-3.49) | 0.732 |
| Graduate/Professional | 5.46 (1.87-15.92) | 0.002 | 1.00 (0.21-4.73) | 0.998 |
| **Age** |  |  |  |  |
| 18-25 | Reference |  | Reference |  |
| 26-35 | 1.56 (0.91-2.69) | 0.108 | 1.45 (0.72-2.94) | 0.298 |
| 36-45 | 0.94 (0.55-1.61) | 0.830 | 0.52 (0.26-1.05) | 0.067 |
| 46-55 | 0.73 (0.42-1.27) | 0.270 | 0.49 (0.24-1.00) | 0.051 |
| 55+ | 1.15 (0.69-1.90) | 0.596 | 0.61 (0.31-1.17) | 0.137 |

CI = Confidence Interval
\*Model was adjusted for all other variables in the table

## Discussion

The low levels of knowledge about Monkeypox and the strong association between COVID-19 vaccination status and attitudes towards a Monkeypox vaccine, indicate the clear need for more communication about the outbreak. In recent outbreaks such as the 2014-2016 Ebola outbreak and the COVID-19 pandemic, lack of early and clear communication created space for misinformation, which in turn had a major impact on the effectiveness of outbreak control measures.^5,6^ Even though currently no recommendation regarding a Monkeypox vaccine for the general public has been made, interventions should leverage trusted sources of information, such as healthcare professionals and officials, but also doctors and researchers with a large online following.

## Data Availability

All data produced in the present study are available upon reasonable request to the authors

