## supplement for "Attitudes of the US general public towards Monkeypox"

### Supplemental material

Survey weights were created based on age, gender and race and sourced from the American Community Survey (<https://www.census.gov/programs-surveys/acs/data.html>). In the table below we show the demographics of the sample weighted (as summarized in the research letter), unweighted and compare it to the American population based on the American Community Survey data from 2020.

|  | Unweighted  sample  N (%) | Weighted  sample  % | American  population  % |
| --- | --- | --- | --- |
| **Gender** |  |  |  |
| Male | 410 (48) | 49 | 49 |
| Female | 436 (51) | 51 | 51 |
| Other | 10 (1) | 0 |  |
| **Age (years)** |  |  |  |
| 18-25 | 103 (12) | 12 | 9 |
| 26-35 | 168 (20) | 18 | 13 |
| 36-45 | 157 (18) | 16 | 12 |
| 46-55 | 150 (18) | 17 | 12 |
| 55+ | 278 (32) | 38 | 33 |
| **Race** |  |  |  |
| Black/African American | 113 (13) | 13 | 12 |
| American Indian/Alaska Native | 17 (2) | 2 | 1 |
| Asian | 39 (5) | 4 | 6 |
| Native Hawaiian/Other Pacific Islander | 2 (0) | 0 | 0 |
| White | 685 (80) | 81 | 62 |
| **Ethnicity** |  |  |  |
| Hispanic | 82 (10) | 9 | 19 |
| Non-hispanic | 774 (90) | 91 | 82 |
| **Education** |  |  |  |
| No high school | 27 (3) | 3 | 12 |
| High school | 248 (29) | 28 | 27 |
| Some college | 233 (27) | 28 | 20 |
| College | 230 (27) | 28 | 29 |
| Graduate/Professional | 118 (14) | 13 | 13 |
